# Reducing squalene epoxidase by the aging-dependent intra-tissue cholesterol accumulation is associated with increased colorectal cancer patient severity in high-risk populations

**DOI:** 10.1101/2023.03.28.23287791

**Authors:** Soo Young Jun, Hyang Ran Yoon, Ji-Yong Yoon, Jeong-Ju Lee, Jin-Man Kim, Nam-Soon Kim

## Abstract

**Objective:** Recently, we demonstrated cholesterol accelerating colorectal cancer (CRC) progression via squalene epoxidase (SQLE) reduction, activating the β-catenin oncogenic pathway while downregulating the p53 pathway, mediated by the inhibition of GSK3β activity (GSK3β^pS9^). However, the interrelationship between cholesterol increase and CRC progression with aging has never been determined.

**Design:** We utilized case data from public databases and human specimens to assess the relationship between cholesterol accumulation and CRC progression with aging. Digital image analysis-machine learning with multiplex fluorescence-immunohistochemistry evaluated the effects of SQLE, p53^WT^, p53^MT^, and GSK3β^pS9^ (hereafter candidates) on the survival of CRC patients. Also, the prognostic and diagnostic abilities were assessed by a time-dependent receiver operating characteristic (timeROC) and a ROC curve with and without the discriminant score for the candidates as a single or whole, respectively.

**Results:** We found an accumulation of cholesterol and cholesteryl ester in tissues with aging, which led to the acceleration of CRC progression through substantial decreases of SQLE, p53^WT^, p53^MT^ expressions and inhibition of GSK3β activity in advanced CRCs. Retrospective studies demonstrated that SQLE significantly impacted the shortened progression-free survival of the population with progressive pathological severity and high CRC risk beyond the age of 50. Clinical assays further showed the excellent prognostic and diagnostic abilities of SQLE and GSK3β^pS9^ but also the substantial diagnostic potential of the combined candidate for the aged high-risk CRC population.

**Conclusion:** We provide new insights into the relationship between cholesterol increase and CRC progression with aging and identify valuable biomarkers for aged populations with high-risk CRC.

## INTRODUCTION

Colorectal cancer (CRC) is the second leading cause of cancer-related death globally^1^, even with advances in screening technology and an increasing detection ratio at earlier stages. Moreover, most CRC patients’ deaths are associated with metastatic CRCs^2,3^. Therefore, identifying biomarkers related to patients with high-risk CRC is crucial for implementing appropriate surveillance and treatment protocols.

CRC incidence and mortality increase with age, not only showing significant growth beyond age 50^4^: the most prevalent group at risk of CRC with mostly diagnosed between ages 65 and 74 years, but also a recent noticeable increase in patients under 50 (early-onset CRC). Due to the current rise in high-cholesterol diets, several societies, such as the Korean Society of Gastrointestinal Cancer Research^5^ and the US Preventive Services Task Force^6^, have renewed their CRC screening recommendation to include adults ages 50 to 75 but also those ages 45 to 49 years. Abualkhair’s recent report^7^ further supports that using a subsequent one-year age model, the significant ratio of CRC incidence between ages 49 and 50 represents unidentified early-onset CRC detected later via screening after age 50. Moreover, despite ongoing debate, multiple epidemiological studies point to a cholesterol/low-density lipoprotein-related lifestyle as the cause for increased CRC fatality^8,9^ and early-onset CRC incidence^10^. However, the age-dependent interrelationship between cholesterol increase and CRC progression has never been elucidated.

Nevertheless, of its essential to maintaining cell membrane integrity and providing the precursor for several vital substances such as steroid hormones and vitamin D, overproduction of cholesterol harms the cells but also the organs. Relatedly, studies have reported that high serum cholesterol levels contribute to malignant colonic transformation by causing hypoxia in large intestine tissues^11,12^. Thus, the delicate adjustment of intra-cellular cholesterol content is required. Squalene epoxidase (SQLE) has been reported as a bona fide oncogene in several cancers, including breast^13^ and liver^14^, and also is considered a rate-limiting enzyme in cholesterol biosynthesis^15,16^. In addition, Gill et al., and others reported that cholesterol exquisitely regulates the SQLE gene and protein by suppressing SQLE expression and inducing proteasomal degradation of the SQLE protein via MARCHF6-ubiquitination at the N-terminal 100 amino acid^15,16^. Notably, we demonstrated this disconnection under certain pathologic conditions on the expression of the SQLE gene and protein^17^; SQLE degradation caused by accumulated cholesterol over a certain threshold accelerates CRC progression and metastasis via activating the β-catenin oncogenic pathway by inhibiting the p53 anti-tumor suppressor path and GSK3β activity. In this study, we assessed the age-dependent relationship between cholesterol increase and CRC progression using CRC specimens grouped according to patient age and cancer grade with a machine learning analysis of digitized whole slide images through retrospective evaluation. We found the underlying relationship between aging-dependent intra-tissue cholesterol augmentation and CRC progression via SQLE reduction for the old population with high CRC risk beyond age 50.

## MATERIALS AND METHODS

### Sources of patients’ samples

We constructed a discovery cohort, split into training and testing sets, using human specimens supplied by TissueArray.com (previously, US Biomax, Derwood, MD, USA; Table 1). Additionally, we validated the results of this discovery cohort using retrospectively collected human samples from Chungnam National University Hospital (Daejeon, South Korea). To determine the content of intra-tissue total cholesterol and cholesteryl ester, as well as the intra-tissue levels of the candidates, we obtained frozen human CRC samples with patient information from BIOBANK (Chungnam National University Hospital, Daejeon, South Korea). The studies with TissueArry.com and Chungnam Nation University Hospital were exempted by the Public Institution Bioethics Committee designated by the Ministry of Health and Welfare and the Research Ethics Committee of the Korean Research Institute of Bioscience and Biotechnology (P01-202104-31-006 and 2021-0838-001, respectively).

**Table 1.** Clinicopathological characteristics of the discovery cohort. (a), SD, standard deviation.

| Discovery phase | Case-control cohort |  |  |
| --- | --- | --- | --- |
| All patients | 1638 |  |  |
| Cohort size |  | Patients with Cancer | Control subjects |
|  | n (%) | 1311 (80.0%) | 327 (20.0%) |
| Sex |  | Patients with Cancer | Control subjects |
| Male | n (%) | 828 (63.1%) | 201 (61.5%) |
| Female |  | 483 (36.9%) | 126 (38.5%) |
| Age | Overall | Patients with Cancer | Control subjects |
| Mean | 57.1 | 57.4 | 56.3 |
| Median ± SD<sup>(a)</sup> | 58±13.9 | 58±13.5 | 66±15.3 |
| Range (min - max) | 24-86 | 24-86 | 25-82 |
| Subtype |  | Patients with Cancer | Control subjects |
| Adenocarcinoma | n (%) | 1156 (88.2%) | - |
| Mucinous adenocarcinoma |  | 145 (11.1%) | - |
| Papillary carcinoma |  | 9 (0.7%) | - |
| Signet-ring cell carcinoma |  | 1 (0.0%) | - |
| Control |  | - | 327 |
| AJCC stage |  | Patients with Cancer | Control subjects |
| S1 - S2 | n (%) | 471 (35.9%) | - |
| S3 - S4 |  | 800 (61.0%) | - |
| unknown |  | 40 (3.1%) | - |
| Control |  | - | 327 |
| The TNM staging |  | Patients with Cancer | Control subjects |
| T3N0M0 (T1N0M0, T2N0M0) |  | 213 (16.2%) | - |
| T3N1M0 (T3N1M1) |  | 295 (22.5%) | - |
| T3N2M0 (T3N2M1) |  | 193 (14.7%) | - |
| T4N0M0 (T4N0M1) |  | 218 (16.6%) | - |
| T4N1M0 (T4N1M1) |  | 145 (11.1%) | - |
| T4N2M0 (T4N2M1) |  | 158 (12.1%) | - |
| unknown |  | 89 (6.8%) | - |
| Control |  | - | 327 |
| Tumour differentiation |  | Patients with Cancer | Control subjects |
| G1 | n (%) | 396 (30.2%) | - |
| G2 |  | 646 (49.3%) | - |
| G3 |  | 162 (12.4%) | - |
| Control |  | 107 (8.1%) | - |
| Follow up | Overall | Patients with Cancer | Control subjects |
| Mean | 93.6 | 91.7 | 101.9 |
| Median ± SD | 110±43.1 | 109±43.4 | 123±40.4 |
| Range (min - max) | 2-166 | 2-166 | 30-166 |

### The content of intra-tissue total cholesterol and cholesteryl ester

The content of intra-tissue total cholesterol and cholesteryl ester were determined according to the manufacturer’s instructions. Briefly, CRC tissues were homogenized in a mixture (chloroform: isopropanol: NP-40; 7:11:0.1, respectively). Then, every liquid phase after centrifugation at 15,000g was vacuum air-dried at 50°C for 30 minutes. To measure the total cholesterol and free cholesterol, the reaction mixture with and without cholesterol esterase was mixed with the extraction dissolved in the assay buffer. The total and free cholesterol levels were determined at OD570nm after incubating the reaction mixture at 37°C for 60 minutes. The cholesteryl ester concentration was calculated by subtracting the value of the free cholesterol from the total cholesterol value.

### Western blotting

A Western blot was performed, as we previously reported^17^. Briefly, tissue lysates were prepared in ice-cold Pierce IP Lysis/Wash Buffer (Thermo Fisher Scientific, 87788), separated by sodium dodecyl sulfate-polyacrylamide gel electrophoresis, transferred onto nitrocellulose membranes, and blocked with the indicated antibodies also used for multiplex fluorescence-based immunohistochemistry. The specificity of the antibodies was evaluated in our previous study^17^.

### A multiplex fluorescence-based immunohistochemistry

As previously described^17^, the levels of SQLE, p53^WT^, p53^MT^, and GSK3β^pS9^ in human tissues were determined. Briefly, deparaffinized and antigen-retrieved slides were reacted with antibodies against SQLE, p53^WT^, GSK3β (Santa Cruz Biotechnology, Dallas, TX, USA; SC-99144, SC-126, SC-377213, respectively), p53^MT^ (Abcam, Boston, MA, USA; ab32049), and GSK3β^pS9^ (Cell Signaling Technology, Danvers, MA, USA; 9323s) overnight. Afterward, each slide was reacted with Alexa Fluor-633 goat anti-rabbit, −546 goat anti-mouse, or −488 goat anti-mouse antibody (Thermo Fisher Scientific, Waltham, MA, USA; A21071, A11030, or A11001, respectively). Confocal observation (LSM800, Carl Zeiss, Oberkochen, Germany) was then performed. To evaluate antibody specificity, thyroid cancer tissues were stained with the same antibodies and then observed under a confocal microscope (Supplementary Figure 4), confirming the negative responsiveness of the SQLE antibody in thyroid cancer given in the report “The human protein atlas (https://www.proteinatlas.org/ENSG00000188404-SELL/pathology).

### Statistical analysis

An unpaired *t*-test was used to determine a statistical significance; all *p* values were noted in each figure. All statistical analysis was performed using *R* and *SPSS*. For the quantitative analysis of SQLE, p53^WT^, p53^MT^, and GSK3β^pS9^ (hereafter candidates) in CRC tissues, we performed global normalization^18^ as follows; once divided by GSK3β median intensity, each value was log-transformed, then individual log values were subtracted by the median log value of each group. The levels of the candidates were represented as the median with the standard error of the mean. A uni-, multivariate Cox proportional hazard and Kaplan-Meier method based on the log-rank test were used for survival analyses. To determine the prognostic and diagnostic abilities of the candidates, we applied a time-dependent receiver operating characteristic (timeROC) and ROC curves with or without a discriminant score. TimeROC utilized a weighted Cox regression for survival data to determine the area under the timeROC curve. The timeROC and ROC curves without a discriminant score were obtained using *R*, while ROC curves with a discriminant score were based on *SPSS*.

## RESULTS

### Epidemiological study

We used case data from the Surveillance, Epidemiology, and End Results (SEER) program to verify the recent incidence and mortality trend of CRC. Based on the SEER guidelines^19^, we categorized the following cancer sub-types as colon cancer: appendix, ascending, cecum, descending, transverse, sigmoid, hepatic flexure, splenic flexure, and rectosigmoid segment colon, excluding anal cancer^20^ due to most cancerous changes similar to cervical cancer infected with human papillomavirus (HPV).

Our analysis verified the significant statistical ratio between ages 49 and 50 in which early-onset CRC (CRCs diagnosed before 50) was detected beyond age 50 via screening uptake (Supplementary Figure 1A), consistent with Abualkhair’s report^7^. In addition, we evaluated increased early-onset CRC incidence^10^ and mortality (Supplementary Figure 1B, C) and reduced CRC death rate decline^8,9^ (Supplementary Figure 2), which are considered dietary cholesterol intake-related characteristics. Moreover, given the age-dependent CRC incidence and mortality, we determine if cholesterol levels increase with age using data from the National Health and Nutrition Examination Survey (NHNES). Notably, in both sexes, the percentage of those reporting high cholesterol increased with age, especially in 2015-2018 (Supplementary Figure 3A, B). Therefore, our epidemiological analysis strongly supports an aging-dependent association between CRC incidence/mortality and cholesterol increase.

### Evaluation of cholesterol accumulation and CRC progression with aging

In light of our recent report^17^ on cholesterol accumulation and its acceleration of CRC progression through SQLE reduction, which activates the β-catenin oncogenic pathway and inhibits the p53/p21 tumor suppressor pathway, we examined the expression levels of SQLE, p53^WT^, p53^MT^ and the phosphorylation of GSK3β^pS9^ on age-dependent CRC progression using human tissue microarray sections to reduce between-subject variability. Besides, we employed two types of p53 antibodies, p53^WT^ and p53^MT^, which are reported to detect mainly wild-type and mutant p53, respectively.

We found a significant decrease in the expression of SQLE, p53^WT^, and p53^MT^ and increase in GSK3β^pS9^ phosphorylation between ages 50-70 compared with those between 20-30 (Figure 1A, B). These findings were confirmed by a different cohort (Figure 1C, D). In addition, consistent with our previous report, we corroborated a significantly decreased SQLE, p53^WT^, and p53^MT^ expressions and increased GSK3β^pS9^ phosphorylation during CRC progression (Figure 1E). Of note, we verified the significant downregulation of p53^WT^ but also p53^MT^ in the aged and aggressive CRCs, partially explained by our previous demonstration^17^ that overexpression of p53 wild type, but also p53R273H, having partial transactivation activity^17^, with the treatment of HLI373, an Mdm2 inhibitor suppressed cell survival in SQLE-reduced condition. Moreover, the involvement of SQLE, p53^WT^, p53^MT^ and GSK3β^pS9^ during CRC progression was further supported by the staining of thyroid cancer tissues with antibodies against the candidates then observed under confocal microscopy (Supplementary Figure 4), showing no age-dependent difference, especially in SQLE expression and therefore corroborating our previous report that SQLE reduction due to cholesterol accumulated is a distinctive feature during the progression of gastrointestinal cancers.

**Figure 1.**
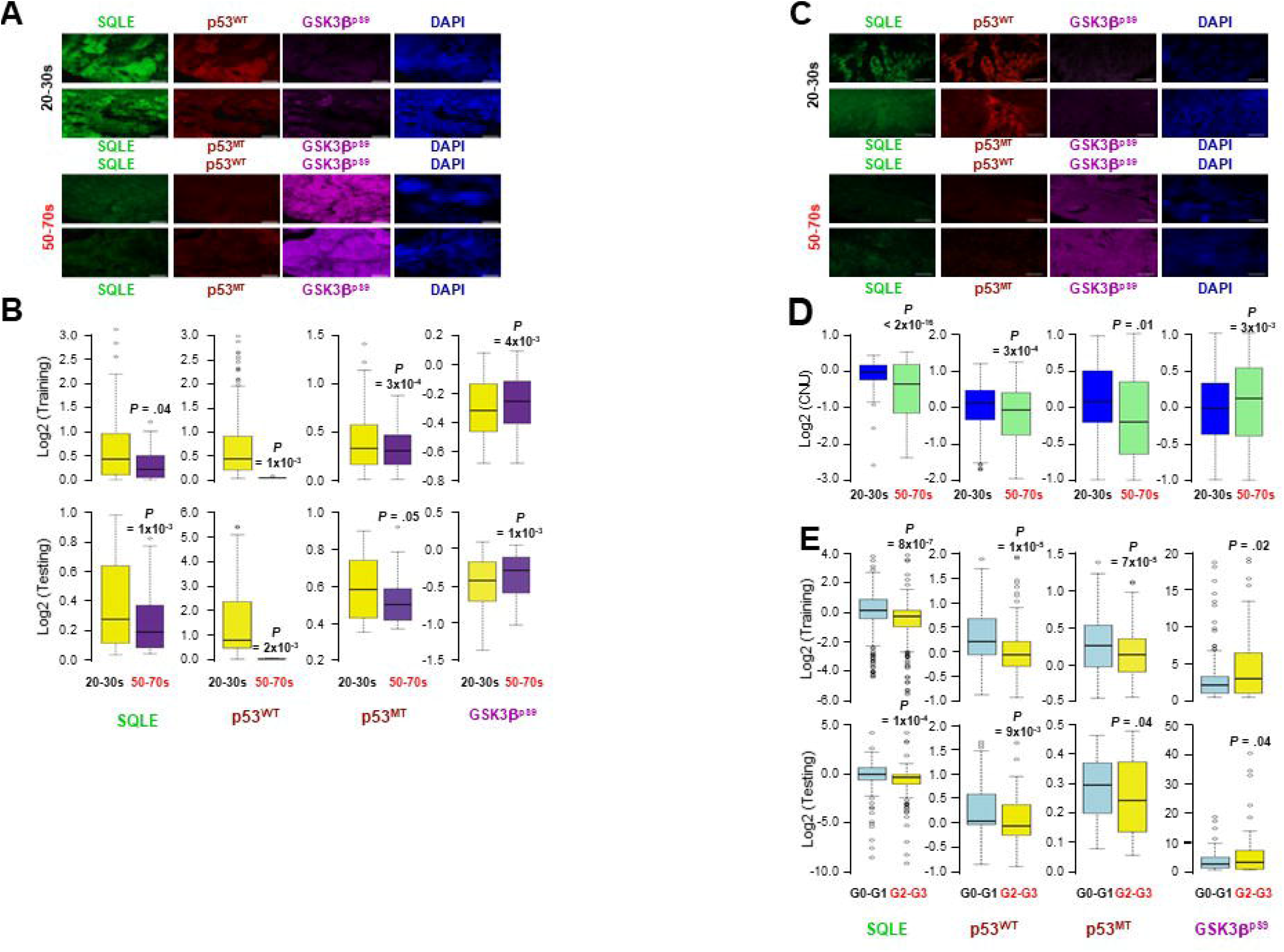
Involvement of SQLE, p53^WT^, p53^MT^, and GSK3β^pS9^ in aging-dependent CRC progression. Each CRC specimen grouped according to patient age (A-D) or CRC grade (E) was stained simultaneously with antibodies against the indicated candidate and then observed under confocal microscopy. The quantification of each variable shown in (A) and (C) was presented in (B) and (D), respectively (Table 1). The tissues were supplied by TissueArray (A-B, E) or Chungnam National University Hospital (C-D). p53^WT^, mainly detecting wild-type p53 (DO-1), p53^MT^; mainly mutant p53 (Y5), and GSK3β^pS9^: the inactive form of GSK3β determined by the anti-GSK3β^pS9^ antibody. Scale bar, 100 μm. *\**An unpaired t-test determined *P* values.

We then assessed the age-dependent relationship between cholesterol augmentation and CRC progression using CRC tissue lysates. The intra-tissue levels of total cholesterol and cholesteryl ester significantly increased between ages 20-30 and 50-70 (Figure 2A, B). Notably, cholesterol failed to show a significant increase between G2 and G3 (Figure 2A, B) as a higher accumulation of cholesterol within G3 tissues degraded SQLE further (Figure 2C: red boxes, D). Moreover, consistent with the observation in the digital analysis using tissue microarray (Figure 1), the augmented reduction of SQLE expression further accelerated CRC progression, as evidenced by a further decrease in p53^WT^ and p53^MT^ expressions and increase in GSK3β^pS9^ phosphorylation in advanced CRCs (Figure 2C: red boxes, D).

**Figure 2.**
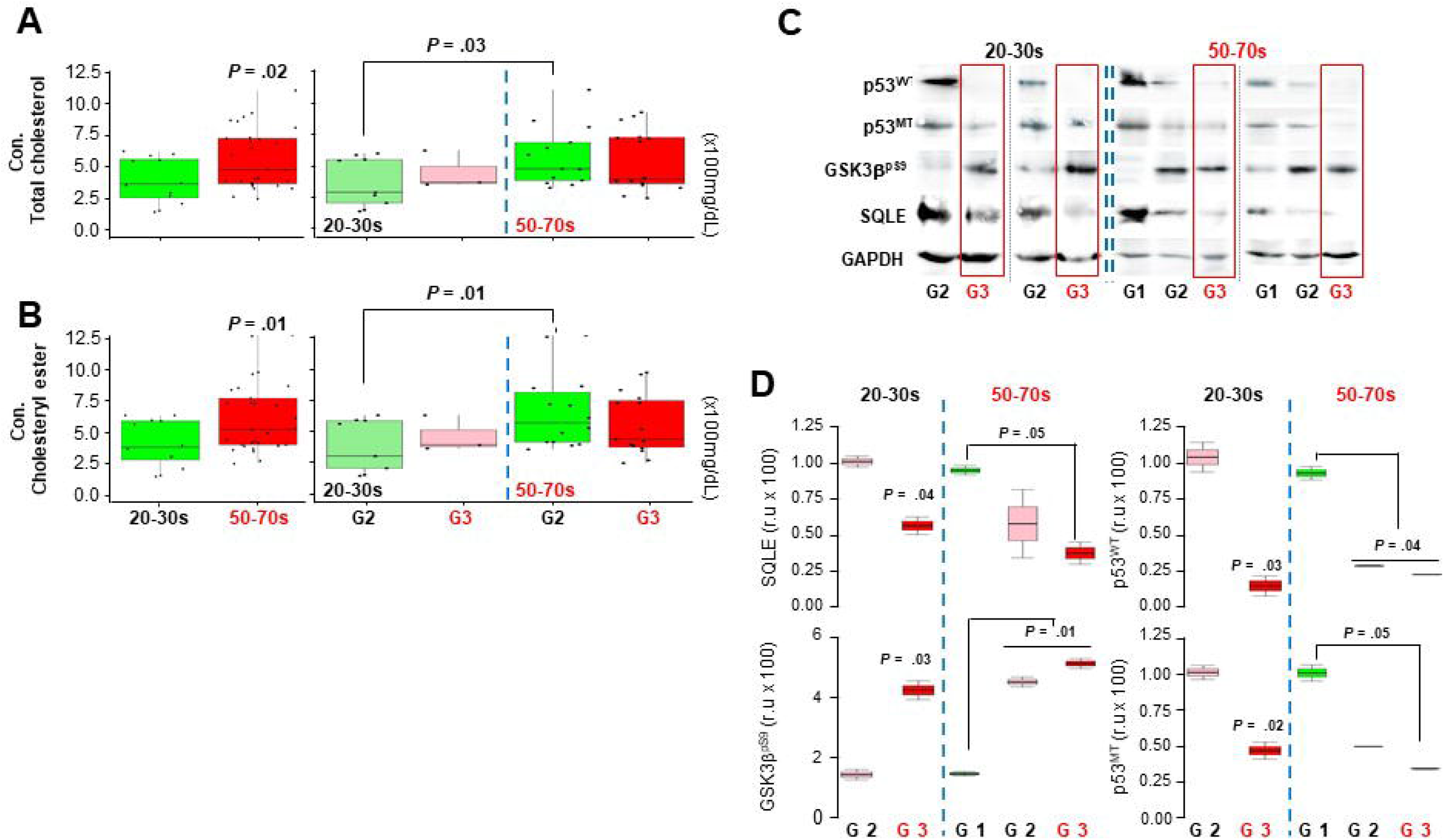
Relevance of cholesterol augmentation in age-related CRC progression. The total cholesterol (A) and cholesteryl ester (B) in CRC tissue lysates were measured according to the manufacturer’s instructions. Dots indicate a single CRC sample. (C) For Western blot analysis, ten independent human CRC specimens were used. For loading control, GAPDH was used. (D) ImageJ (National Institute of Health, Bethesda, Maryland, USA) was used to quantify protein expression, r.u, relative uncertainty, and *P* values were calculated by an unpaired t-test. p53^WT^, mainly detecting wild-type p53 (DO-1); p53^MT^, mainly to detect mutant p53 (Y5); GSK3β^pS9^, the inactive form of GSK3β.

Overall, the results demonstrate that aging-dependent accumulated cholesterol inside tissue expedites CRC progression via additional reduction of SQLE, leading to increased downregulation of p53^WT^ and p53^MT^ expression by upregulated inhibition of GSK3β activity in advanced CRCs.

### Relevance of high-risk CRC candidates

Considering the cholesterol augmentation-CRC aggressiveness axis, we then assessed the potential effects of SQLE, p53^WT^, p53^MT^, and GSK3β^pS9^ (hereafter candidates) on increasing CRC risk through a retrospective analysis of collected specimens from CRC patients and controls. Table 1 shows the clinicopathological characteristics of the discovery cohort; The cohort consisted of twice as many male patients, consistent with most reports on male prevalence in CRC incidence and mortality, as well as adenocarcinoma (88.2%), the most common subtype found in approximately 90% of CRC patients. Also, this cohort has TNM staging, with a similar patient distribution across all stages, enabling estimation of CRC aggressiveness and the cancer patients having shortened progression-free survival (PFS) compared with the control. Therefore, this discovery cohort has sufficient characteristics to evaluate the candidates’ clinical implications in increasing CRC progression and patient survival (Table 1).

Next, we randomly split the discovery cohort into training (70%) and testing sets (30%) by apply a random forest-based modeling based on the machine-learning approach (Table 1; Supplementary Table 1, 2). Then, the Cox proportional hazard regression analysis (hereafter Cox analysis) was used to determine whether pathological-related variables and the candidates as a single or whole impact CRC patients’ PFS. Notably, we found the significant effects of not only Sex (males), the pathological variables including stages, TNM, and grade, but also SQLE on the survival of CRC patients beyond 50 (Supplementary Tables 1, 2; Figure 3). Interestingly, while p53^WT^ and GSK3β^pS9^ showed a significant hazard ratio (HR) associated with the survival of the aged population in the testing and training sets, respectively: this HR ratio was not confirmed in the other set, possibly caused by the bias of censored data to one set. Furthermore, compared with patients under 50, the pathological-related variables, including stage, TNM, and grade, showed a significantly increased HR ratio regarding the survival of CRC patients beyond 50, indicating increased pathological severity in the aged CRC patients (Figure 3; Supplementary Tables 1, 2). Moreover, the increased pathological severity observed in older CRC patients was consistent was a shortened PFS for CRC patients beyond 50 compared to those under 50, as demonstrated by the Cox and KM analyses (Figure 3, 4; Supplementary Tables 1, 2).

**Figure 3.**
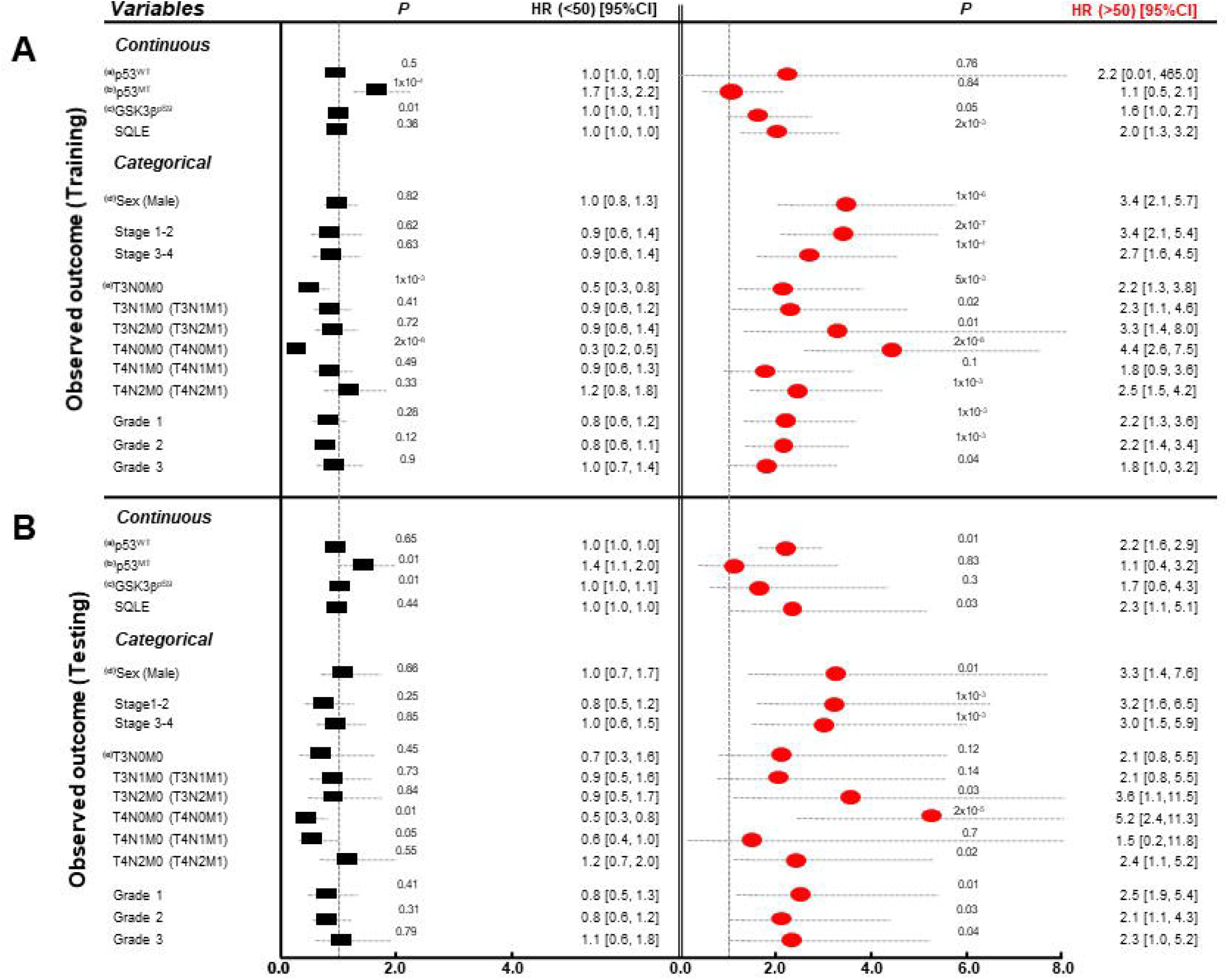
Multivariate analysis of the candidates and pathological factors related to survival in an independent set generated from the discovery cohort by applying random forest and then separated by before and after age 50. (a) p53^WT^: wild-type p53 (DO-1), (b) p53^MT^: mutant p53 (Y5), (c) GSK3β^pS9^: the inactive form of GSK3β (the anti-GSK3β^pS9^ antibody), (d) Sex (Male): risk assessment for males compared to females, and (e) T3N0M0 including T1N0M0 and T2N0M0. *P* values are noted on the dashed line of HR weights. (A) Training and (B) testing sets.

**Figure 4.**
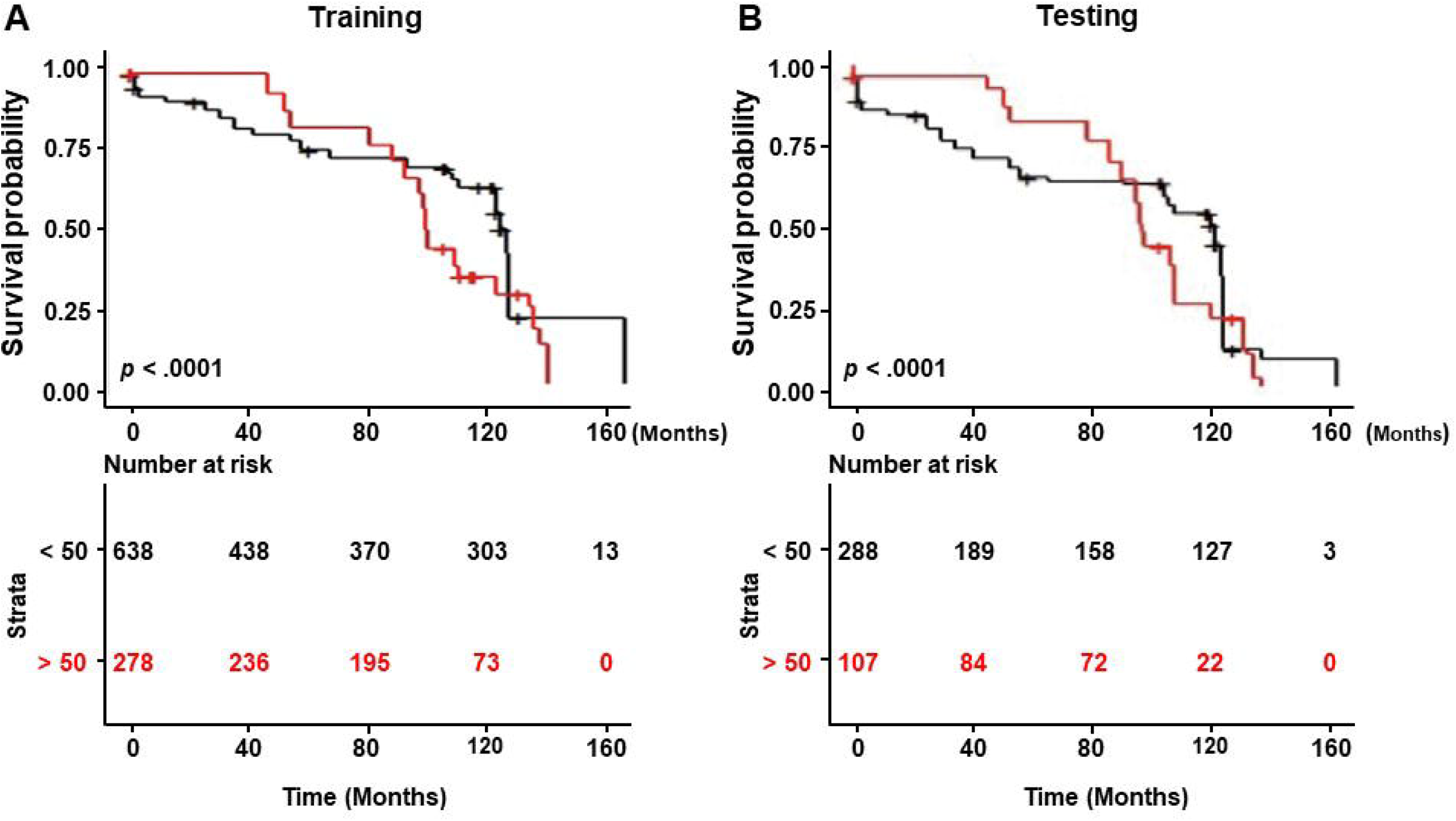
Overall survival analysis using a Kaplan-Meier plot for an independent set. Each set was split into before and after age 50, and a Log-rank *p*-test was used. The median survival months and 95% CIs for the population under and over the age of 50 in the training and test sets were 124 (124-126), 99 (98-100), 124 (110-126), and 99 (97-126), respectively.

Overall, our study provides evidence for the crucial role of SQLE on the survival of the aged populations with a high risk of CRC showing increased pathological severity, in contrast to p53^MT^ impacting the survival of those under 50 (Figure 3, 4; Supplementary Table 1, 2).

### Clinical verification of candidates in populations at high CRC risk

We further evaluated whether the candidates could distinguish between patients who would die or survive within a given time (t). For this purpose, we utilized a time-dependent ROC (timeROC)^21^ analysis that employed a weighted Cox regression for survival data to determine the area under the timeROC curve: the times that maximize the area under the curve (AUC) ratio was 123 and 137 months for patients before and after 50, respectively (Figure 5). Importantly, SQLE, which showed a critical impact on the PFS of the aged population with high CRC risk (Figure 3), also demonstrated remarkable prognostic ability in discriminating between CRC patients over 50 who would die or survive within 137 months but not in those under 50 (Figure 5). Interestingly, although the significant HR values of GSK3β^pS9^ regarding the survival of the aged populations at the training test failed to be confirmed in the testing set, we found that, unlike p53^WT^, which showed its significant association with the aged populations in the testing set, GSK3β^pS9^ had an important prognostic ability verified in both sets (Figure 5).

**Figure 5.**
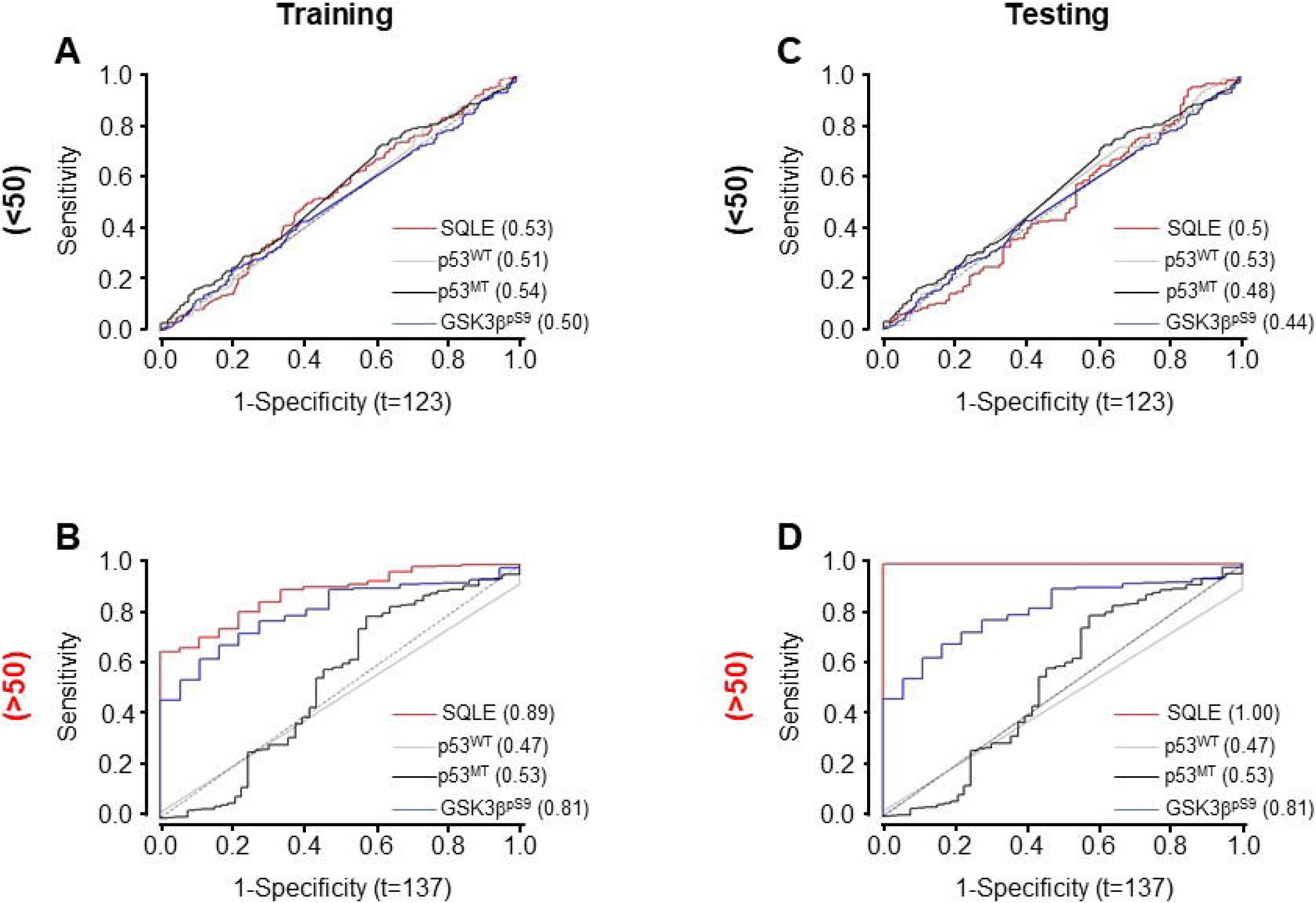
Time-dependent ROC analysis for prognostic patient survival within a given time. The time was decided to maximize the ROC curve utilizing a weighted Cox regression. (A-B) Training and (C-D) testing sets, and the times for patients before (A, C) and after (B, D) age 50 were 123 and 137, respectively. p53^WT^, mainly detecting wild-type p53 (DO-1), p53^MT^; to detect mutant p53 (Y5), and GSK3β^pS9^: the inactive form of GSK3β (the anti-GSK3β^pS9^ antibody).

Consistent with the prognostic abilities, SQLE and GSK3β^pS9^ showed satisfactory diagnostic efficiencies with AUC values of 0.8 and 0.7 in distinguishing CRCs from controls, respectively, in contrast to p53^WT^ and p53^MT^ (Supplementary Figure 5). Moreover, the excellent diagnostic capacities of SQLE and GSK3β^pS9^ were corroborated by the ROC curve, which utilized the discriminant score derived from each candidate in distinguishing CRC patients from controls (Figure 6A, B). Notably, when all candidates were merged to calculate the discriminant score for CRC patients above 50, a significant differential distribution was observed compared to the control group. However, this was not observed in patients under age 50 (Figure 6C-F: left). The observed differential distribution conformed with the excellent AUC ratios of 0.87 and 0.85 in patients over 50 in the training and testing sets, respectively, compared to AUC values of 0.48 and 0.65 in patients under 50 (Figure 6C-F: right).

**Figure 6.**
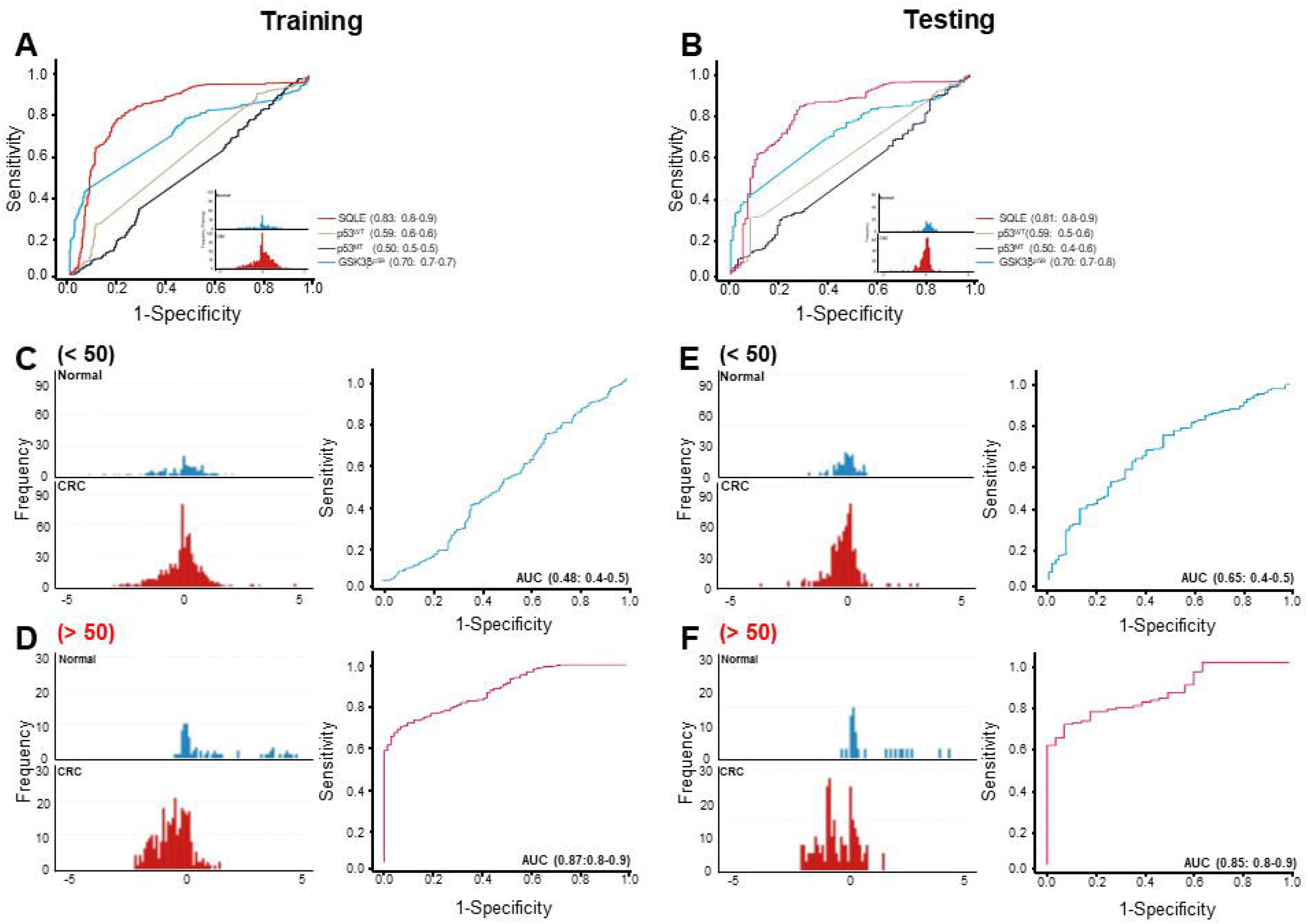
Diagnostic analysis using the discriminant scores for each candidate as a single or whole. The receiver operating characteristic (ROC) curves of the discriminant scores for each candidate in the (A) training and (B) testing sets are shown. (C-F) Each set is divided into before (C, E) and after (D, F) age 50; a linear discriminant distribution for merged candidates (left) and the ROC analysis using the discriminant score for the merged candidates (right) for diagnosing CRC are represented. The ratio of AUC and the corresponding 95% CI are noted. p53^WT^; wild-type p53 (DO-1), p53^MT^; mutant p53 (Y5), and GSK3β^pS9^: the inactive form of GSK3β determined by the anti-GSK3β^pS9^ antibody. (A, C, D) Training and (B, E, F) testing sets.

Therefore, our analysis suggests SQLE and GSK3β^pS9^ as potential biomarkers for prognosis and diagnosis in the aged populations with high-risk CRC. Of note, we also show the substantial diagnostic ability of the merged candidate, SQLE, p53^WT^, p53^MT^, and GSK3β^pS9^ for the aged high-risk CRC population (Figure 5, 6; Supplementary Figure 5).

## Discussion

South Korea, exhibiting an increasing trend in intake of a westernized/cholesterol diet^22^ similar to most developed countries, shows the third-highest global CRC mortality ratio for patients over 50 and the second-highest CRC incidence ratio for those under 50^1^. In this study, we also verified a substantial increase in early-onset CRC incidence and mortality, as well as an age-dependent cholesterol increase, by analyzing case data from the Surveillance, Epidemiology, and End Results (SEER) program and the National Health and Nutrition Examination Survey (NHNES), respectively. However, although its correlation has been suggested^6-8^, the mechanisms underlying cholesterol increase and CRC progression with aging have yet to be fully elucidated.

Recently, we demonstrated^17^ that cholesterol accumulated over a threshold point accelerates SQLE degradation leading to the activation of the β-catenin oncogenic path, mediated by the inhibition of GSK3β activity and the disruption of the p53 tumor suppressor pathway and consequently aggravating CRC progression and metastasis. However, even though our proof-of-concept results show the cause-and-effect relationship between cholesterol increase and CRC progression, our conclusion needed to be revised because most of our studies were performed in cell lines and animals. Therefore, we applied human CRC and control specimens grouped according to patient age and cancer grade, as well as multiplex fluorescence immunohistochemistry with digital image analysis based on machine learning to evaluate the age-dependence between cholesterol increase and CRC progression.

Despite reports linking obesity and CRC^23^, epidemiological findings between serum lipids, especially in LDL, cholesterol, and CRC risk and progression have been conflicting^24^. In contrast, some studies, including Dessi^25^ et al. and others^26,27,^ have reported a higher total cholesterol level in cancer tissues, including gastrointestinal cancer, than in normal tissues. We also have demonstrated^17^ that the accumulation of cholesterol within tumor cells worsens CRC. This cholesterol buildup is partially caused by the increased expression of the low-density lipoprotein receptor (LDLR) and the decreased expression of Adenosine triphosphate (ATP)–binding cassette transporter 1 (ABCA1) in gastrointestinal cancers. As an extension of our recent report, we found a significant augmentation of the intra-tissue total cholesterol and cholesteryl ester with aging accelerating CRC progression; the more cholesterol accumulated in aged CRC tissues, the more SQLE degraded, resulting in an increased lower expression of p53^WT^ and p53^MT^ and a higher level of GSK3β^pS9^ in advanced CRCs. Therefore, our study provides the underlying mechanisms between the aging-dependent intra-tissue cholesterol accumulation and CRC progression via SQLE reduction.

Despite technological advances, the five-year survival rate for distant colorectal cancer is only 17%, compared to 90% for localized cancer^28^. Moreover, CRC mortality has increased from fourth place in 2018 to third place globally in 2020, despite most cancers showing a decline in mortality rates, according to Global Cancer Observatory (GLOBOCAN), together, suggesting new biomarkers are required for populations with high CRC risk. In this study, we demonstrated a critical association between SQLE, controlled by cholesterol, and the shortened progression-free survival (PFS) of populations with high-risk CRC beyond age 50, having increased pathological severity, in contrast to p53^MT^ in those under 50. Besides, although it failed to be confirmed in both sets, which may be caused by the bias of censored patient survival, we found that p53^WT^ and GSK3β^pS9^ had a statistical association with the aged populations in testing and training sets, respectively. Furthermore, clinical assays showed the excellent prognostic and diagnostic capabilities of SQLE but also GSK3β^pS9^ for the aged populations with high-CRC risk beyond age 50. Interestingly, the discriminant assays using the merged candidates showed substantial diagnostic ability in distinguishing from control to CRC for the aged high CRC populations. Therefore, our study provides novel biomarkers including SQLE, GSK3β^pS9^, and the merged candidates for the aged high-risk CRC population to implement appropriate surveillance and treatment protocols.

Previously, we demonstrated^17^ that overexpression of ectopic p53 wild type and p53R273H, which had a partial transactivation activity, along with the treatment of HLI373, an Mdm2 inhibitor, suppressed the survival of SQLE-depleted CRC cells. However, p53-R273H-mTAD, which mutated at four key residues (L22Q/W23SW53W/F54S), which had no functional activity, failed to suppress SQLE-depleted cancer cells’ survival. Thus, in this study, we applied two types of antibodies to detect mainly wild-type (DO-1) and mutant p53, denoted as p53^WT^ and p53^MT^, respectively. We found a statistical reduction of p53^WT^ and p53^MT^ in aggressive and aged CRCs, with p53^WT^ exhibiting approximately twice the relative decrease of p53^MT^. In addition, p53^WT^ had a partial association with the older population in the testing set, while p53^MT^ had a critical association with the survival of the population under 50, confirmed in both sets. However, unlike SQLE and GSK3β^pS9^, which had an association with the survival of the aged population and showed significant diagnostic and prognostic abilities, p53 did not exhibit its efficacy, likely because p53 is a downstream target controlled by SQLE and GSK3β.

Overall, this study corroborates our earlier demonstration that SQLE, via its N-terminal 100 amino acids, directly interacts with active GSK3β and p53, thereby modulating cancer cell proliferation. Conversely, cholesterol helps cancer cells overcome this cellular barrier by reducing SQLE, thereby downregulating the p53/p21 tumor suppressor pathway leading to the activation of the β-catenin oncogenic pathway via GSK3β inhibition. Therefore, these findings underscore the critical role of SQLE, GSK3β^pS9^, and p53 in the survival of aging-dependent accumulated cholesterol-accelerated CRC patients.

Previously, we demonstrated^17^ that a 2% high cholesterol diet induced the pronounced pathological progression and metastasis of CRC by SQLE reduction and robust production of colon-cancer-originating migrating cancer stem cells (collectively indicated by the enrichment of CDCP1 and CD110). Furthermore, in the 2% cholesterol CRC metastasis model, the expression of SQLE was notably decreased in circulating tumor cells (CTCs) extracted using anti-CDCP1 and CD110 antibodies. Thus, in the extension of the current study, we are currently evaluating the diagnostic and prognostic potential of the candidates SQLE, p53^WT^, p53^MT^, GSK3β^pS9^, CDCP1, and CD110 in CTCs.

Despite these promising results, our study has several limitations.

First, we obtained human tissue samples primarily from a commercial company rather than through accredited hospitals. Thus, we tried to avoid possible issues by performing the followings: We prepared a report using the checklist provided by the STrengthening the Reporting of Observational Studies in Epidemiology (STROBE)^29^. In addition, to ensure unbiased data analysis, the discovery cohort was split into a training and testing set using a random forest modeling program which can eliminate the researcher’s bias. Also, the results obtained from the training set were verified by using the testing set and a separate cohort from Chungnam National University Hospital.

Second, we did not investigate the serum levels of lipids or cholesterol in CRC patients evaluated for the intra-tissue levels of candidates and cholesterol accumulation. However, previous epidemiological studies^24^ have shown inconsistent findings regarding the association between serum lipids/cholesterol and CRC risk/progression, unlike other reports, including we^17^ and Dessi^25^, which have consistently demonstrated the accumulation of cholesterol in gastrointestinal CRC tissues and its association with CRC aggressiveness. Our current study further supports the crucial role of intra-tissue accumulated cholesterol and cholesteryl ester in promoting CRC progression.

Third, SQLE has been reported as a bona fide oncogene in several types of cancers^13,14^. Recently, Li C et al., demonstrated^30^ that overexpression of SQLE accelerates the development of colorectal cancer by enhancing cancer cell proliferation and causing gut dysbiosis. In line with this observation, we discovered^17^ that SQLE is upregulated in human CRC compared to normal tissues. Conversely, we and public databases, including Oncomine databases-Simith and Kiaser, found SQLE decrease according to the course of CRC progression, a unique characteristic in gastrointestinal cancers. Therefore, these results imply that SQLE has a dual role in CRC formation and progression, respectively; SQLE contributes to providing the cholesterol necessary for the formation of CRC; on the other hand, cholesterol accumulated inside the tissue downregulated SQLE, thereby accelerating the β-catenin oncogenic pathway through the inhibition of the p53/p21 tumor suppressor pathway and GSK3β activity.

Finally, our study was limited by its small sample size. Thus, a larger cohort study under the supervision of accredited hospitals will be necessary to validate and determine the possible clinical usability of the candidate biomarkers. Finally, our study was limited by its small sample size, and thus a larger cohort study under the supervision of accredited hospitals will be necessary to validate and determine the possible clinical usability of the candidate biomarkers.

In conclusion, our study demonstrates cholesterol accumulation within gastrointestinal tissues during aging and how this process accelerates CRC progression. Furthermore, we have provided novel biomarkers or panels that may be useful in identifying individuals in older populations at high risk for colorectal cancer.

## Data Availability

All data produced in the present work are contained in the manuscript.

## *Abbreviations

SQLE: squalene epoxidase
CRC: colorectal cancer
GSK3β: glycogen synthase 3β
ROC: receiver operating characteristic
timeROC: time-dependent ROC
PFS: progression-free survival
SEER: Surveillance, Epidemiology, and End Results
NHNES: National Health and Nutrition Examination Survey
AUC: area under the curve

## *Acknowledgements

The authors wish to sincerely thank the following people: Dr. Andrew John Brown (University of New South Wales) and Dr. Ngee Kiat Chua (The Walter and Eliza Hall Institute of Medical Research) for advising our study and draft. Professor Austin and Courtney Givens, RN, BSN (Korea Advanced Institute of Science and Technology) for assistance in the preparation of this manuscript.

## REFERENCES

1 https://gco.iarc.fr/today/online-analysis-pie?v=2020&mode=cancer&mode_population=continents&population=900&populations=900&key=total&sex=0&cancer=39&type=1&statistic=5&prevalence=0&population_group=0&ages_group%5B%5D=0&ages_group%5B%5D=17&nb_items=7&group_cancer=1&include_nmsc=1&include_nmsc_other=1&half_pie=0&donut=0

2 https://www.cancer.org/cancer/colon-rectal-cancer/detection-diagnosis-staging/survival-rates.html

3 Wang J, Li S, Liu Y, Zhang C, Li H, et al. Metastatic patterns and survival outcomes in patients with stage IV colon cancer: A population-based analysis. Cancer Med 2020;9:361–373.

4 Colorectal Cancer Facts & Figures 2020-2022, American Cancer Society

5 Choi YJ, Lee DH, Han KD, Kim HS, Yoon H, et al. Optimal starting age for colorectal cancer screening in an era of increased metabolic unhealthiness: a nationwide Korean cross-sectional study. Gut Liver 2018;12:655–663.

6 https://www.uspreventiveservicestaskforce.org/uspstf/recommendation/colorectal-cancer-screening

7 Abualkhair WH, Zhou M, Ahnen D, Yu Q, Wu XC, et al. Trends in incidence of early-onset colorectal cancer in the united states among those approaching screening age. JAMA Netw Open 2020;3:e1920407.

8 Järvinen R, Knekt P, Hakulinen, T, Rissanen, H, Heliövaara M. Dietary fat, cholesterol and colorectal cancer in a prospective study. Br J Cancer 2001;85:357–361.

9 Murphy N, Moreno V, Hughes DJ, Vodicka L, Vodicka P, et al. Lifestyle and dietary environmental factors in colorectal cancer susceptibility. Mol Aspects Med 2019;69:2-

10 Carroll KL, Fruge AD, Heslin MJ, Lipke EA, Greene MW. Diet as a risk factor for early-onset colorectal adenoma and carcinoma: a systematic review. Front Nutr 2022;9:896330.

11 Paraf F, Bouillot JC, Bruneval P. Cholesterol crystal embolisms in adenomatous polyposis coli. Ann Pathol 1999;19:135–138.

12 Herbey II, Ivankova NV, Katkori VR, Mamaeva OA. Colorectal cancer and hypercholesterolemia: review of current research. Exp Oncol 2005;27:166–178.

13 Brown DN, Caffa I, Cirmena G, Piras D, Garuti A, et al. Squalene epoxidase is a bona fide oncogene by amplification with clinical relevance in breast cancer. Sci Rep 2016;6:19435.

14 Liu D, Wong CC, Fu Li, Chen H, Zhao L, et al. Squalene epoxidase drives NAFLD-induced hepatocellular carcinoma and is a pharmaceutical target. Sci Transl Med 2018;10:eaap9840

15 Gill S, Stevenson J, Kristiana I, Andrew JB. Cholesterol-dependent degradation of squalene monooxygenase, a control point in cholesterol synthesis beyond HMG-CoA reductase. Cell Metab 2011;13:260–273.

16 Chua NK, Coates HW, Andrew JB. Squalene monooxygenase: a journey to the heart of cholesterol synthesis. Prog Lipid Res 2020; 79:101033.

17 Jun SY, Andrew JB, Chua NK, Yoon JY, Lee JJ, et al. Reduction of squalene epoxidase by cholesterol accumulation accelerates colorectal cancer progression and metastasis. Gastroenterology 2021;160:1194–1207.

18 Idikio HA. Quantitative analysis of p53 expression in human normal and cancer tissue microarray with global normalization method. Int J Clin Exp Pathol 2011;4:505–512.

19 https://training.seer.cancer.gov/colorectal/anatomy/

20 Stier EA, Chiao EY. Anal cancer and anal cancer precursors in women with a history of HPV-related dysplasia and cancer. Semin Colon Rectal Surg 2017;28:97–101.

21 Kamarudin AN, Cox T, Kolamunnage-Dona R. Time-dependent ROC curve analysis in medical research: current methods and applications. BMC Med res Methodol 2017;17:53.

22 Song SJ, Shim JE, Song WO. Trends in total fat and fatty acid intakes and chronic health conditions in Korean adults over 2007-2015. Public Health Nutr 2019;22:1341–1350.

23 Clinton SK, Giovannucci EL, Hursting SD. The world cancer research fund/American institute for cancer research third expert report on diet, nutrition, physical activity, and cancer: impact and future directions. J Nutr 2020;150:663–671.

24 Fang Z, He M, Song M. Serum lipid profiles and risk of colorectal cancer: a prospective cohort study in the UK biobank. Br J Cancer 2021;124:663–670.

25 Dessi S, Batetta B, Pulisci D, Spano O, Anchisi C, et al. Cholesterol content in tumor tissues is inversely associated with high-density lipoprotein cholesterol in serum in patients with gastrointestinal cancer. Cancer 1994;73:253:258.

26 Eggens I, Ekström TJ, Aberg F. Studies on the biosynthesis of polyisoprenols, cholesterol and ubiquinone in highly differentiated human hepatomas. J Exp Pathol 1990;71:219–232.

27 Liu Y, Guo X, Wu L, Yang M, Li Z, et al. Lipid rafts promote liver cancer cell proliferation and migration by upregulation of TLR7 expression. Oncotarget 2016;7:63856–63869.

28 https://www.cancer.org/cancer/colon-rectal-cancer/detection-diagnosis-staging/survival-rates.html

29 https://www.strobe-statement.org/

30 Li C., Wang Y., Liu D., Wong C.C., Coker O.O., et al., Squalene epoxidase drives cancer cell proliferation and promotes gut dysbiosis to accelerate colorectal carcinogenesis. Gut 2022;71:2253–2265.

