## Supplementary legends for "Reducing squalene epoxidase by the aging-dependent intra-tissue cholesterol accumulation is associated with increased colorectal cancer patient severity in high-risk populations"

**Supplementary Figure 1.** CRC risk and mortality assessment.

**Supplementary Figure 2.** A notable recession in the decrease of CRC fatality rate.

**Supplemental Figure 3.** Assessment of population percentage with hypercholesterolemia according to age.

**Supplemental Figure 4.** The expression levels of the candidates in thyroid cancers.

**Supplementary Figure 5.** The ROC curves for diagnosing CRCs using levels of each candidate.

**Supplementary Table 1.** Clinicopathological information of a training set generated from the discovery cohort by applying random forest and univariate analysis of variables associated with the survival of CRC patients.

**Supplementary Table 2.** Clinicopathological information of the testing set generated from the discovery cohort by applying random forest and univariate analysis of variables associated with the survival of CRC patients.

**SUPPLEMENTARY FIGURE LEGENDS**

**Supplementary Figure 1.** CRC risk and mortality assessment. (A) The one-year age increment of CRC incidence ratio per 100,000 people among ages 40 to 60 (SEER 17 Registries, 2000-2019). (Red) The numbers indicate the difference in CRC incidence ratio between the ages of 49 and 50. CRC Incidence (B) and mortality (C) between patients before and after age 50 (SEER 9 Registries). Incidence rate = (New cancers / Population) x 100,000; Mortality rate = (Cancer deaths / Population) x 100,000. The 2000 US standard population was used for the age adjustment of rates. Incidence rates are adjusted for reporting delays. Source: Incidence – SEER program, 2022. Mortality – NCHS, 2022.

**Supplementary Figure 2.** A notable recession in the decrease of CRC fatality rate. We used data from the SEER cancer statistics review for 1975-2000 (A) and Cancer Stat Facts: Colorectal Cancer (seer.cancer.gov/statfacts/html/colorect.html) for 2001-2020 (B). The numbers (lower left) indicate the median mortality of CRC patients.

**Supplemental Figure 3.** Assessment of population percentage with hypercholesterolemia according to age. The following years were unmarked: 1999-2002, 2003-2006, 2007-2010, and 2011-2014.

Notes: Hypercholesterolemia, also called high total cholesterol, is 240 mg/dL or more. All estimates were age-adjusted by the direct method to the projected 2000 US Census population using the age groups of 20-39, 40-59, and 60 and over. Source: NCHS, National Health and Nutrition Examination Survey, 2015-2018.

**Supplemental Figure 4.** The expression levels of the candidates in thyroid cancers. Thyroid cancer tissues provided by Chungnam National University hospital were stained with antibodies against the candidates and then observed under confocal microscopy. $p53^{WT}$; wild-type p53 (DO-1), $p53^{MT}$; mutant p53 (Y5), $GSK3\beta^{pS9}$: the inactive form of GSK3β (the anti-$GSK3\beta^{pS9}$ antibody). DAPI was used for nucleus staining. Scale bars, 100 μm.

**Supplementary Figure 5.** The ROC curves for diagnosing CRCs using levels of each candidate. The ratio of AUC and the corresponding 95% CI are noted. $p53^{WT}$; wild-type p53 (DO-1), $p53^{MT}$; mutant p53 (Y5), and $GSK3\beta^{pS9}$: the inactive form of GSK3β (the anti-$GSK3\beta^{pS9}$ antibody). (A) Training and (B) testing sets.
