## Supplementary Figures for "Reducing squalene epoxidase by the aging-dependent intra-tissue cholesterol accumulation is associated with increased colorectal cancer patient severity in high-risk populations"

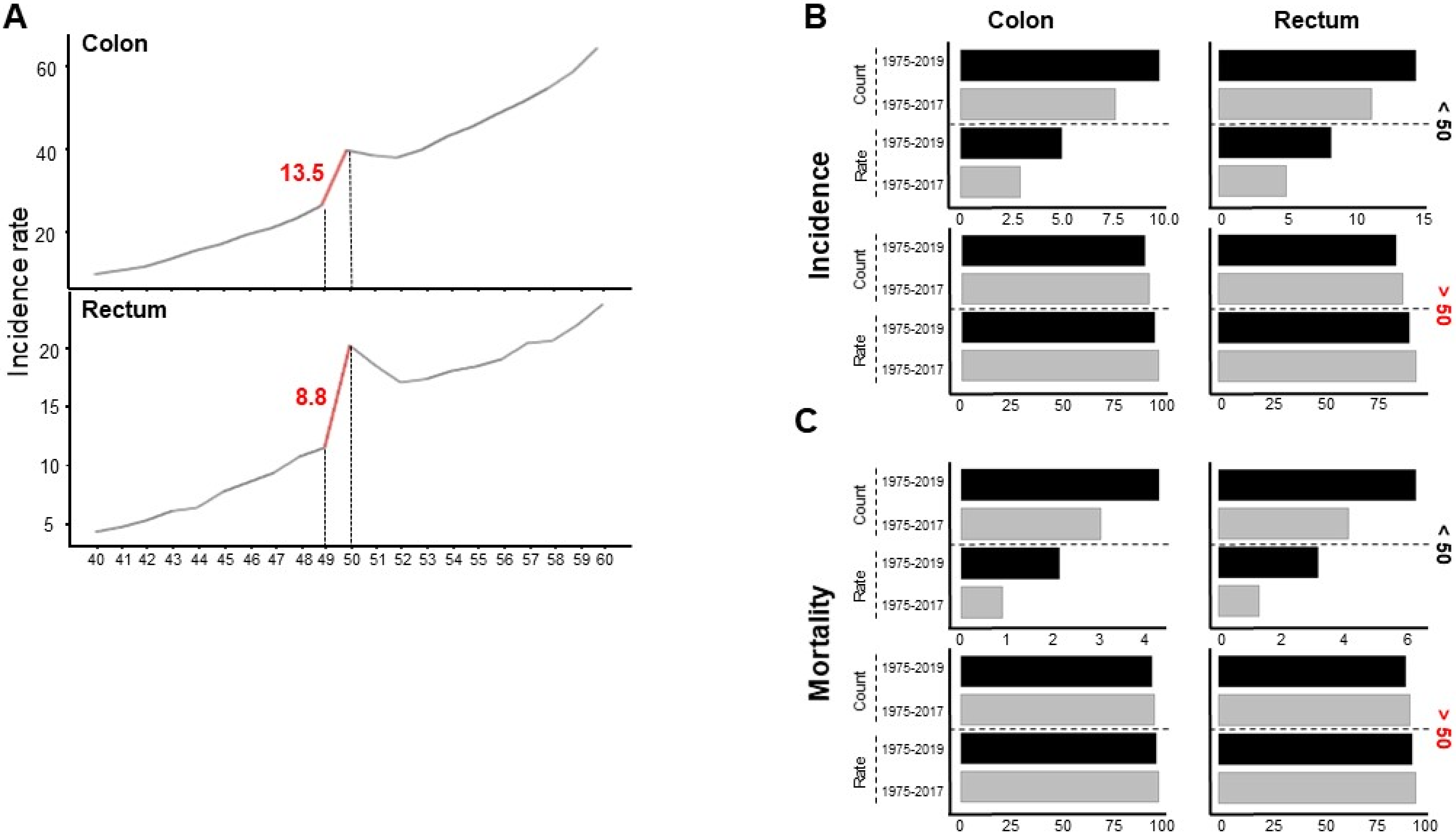


**Supplementary Figure 1. CRC risk and mortality assessment**

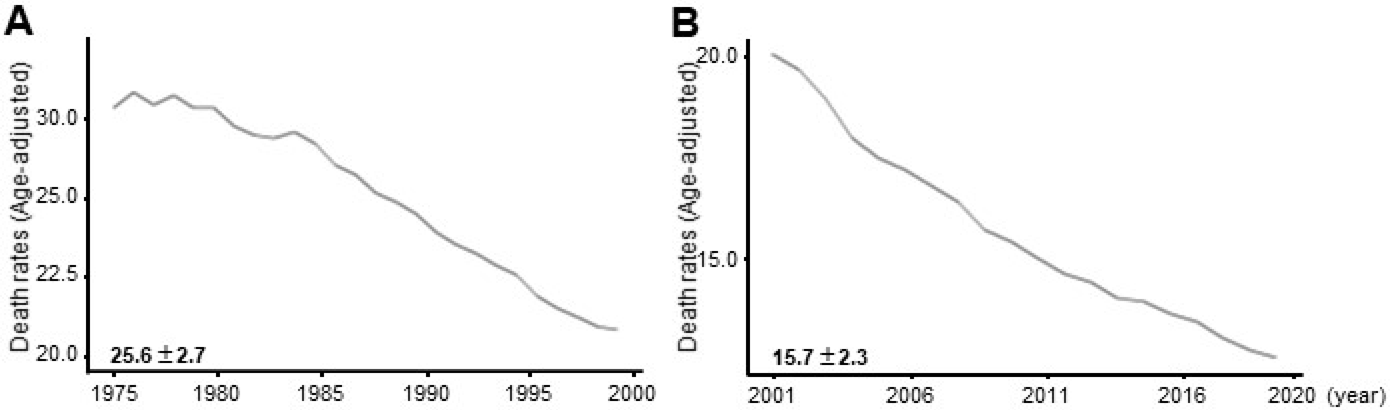


**Supplementary Figure 2. A notable recession in the decrease of CRC fatality rate.**

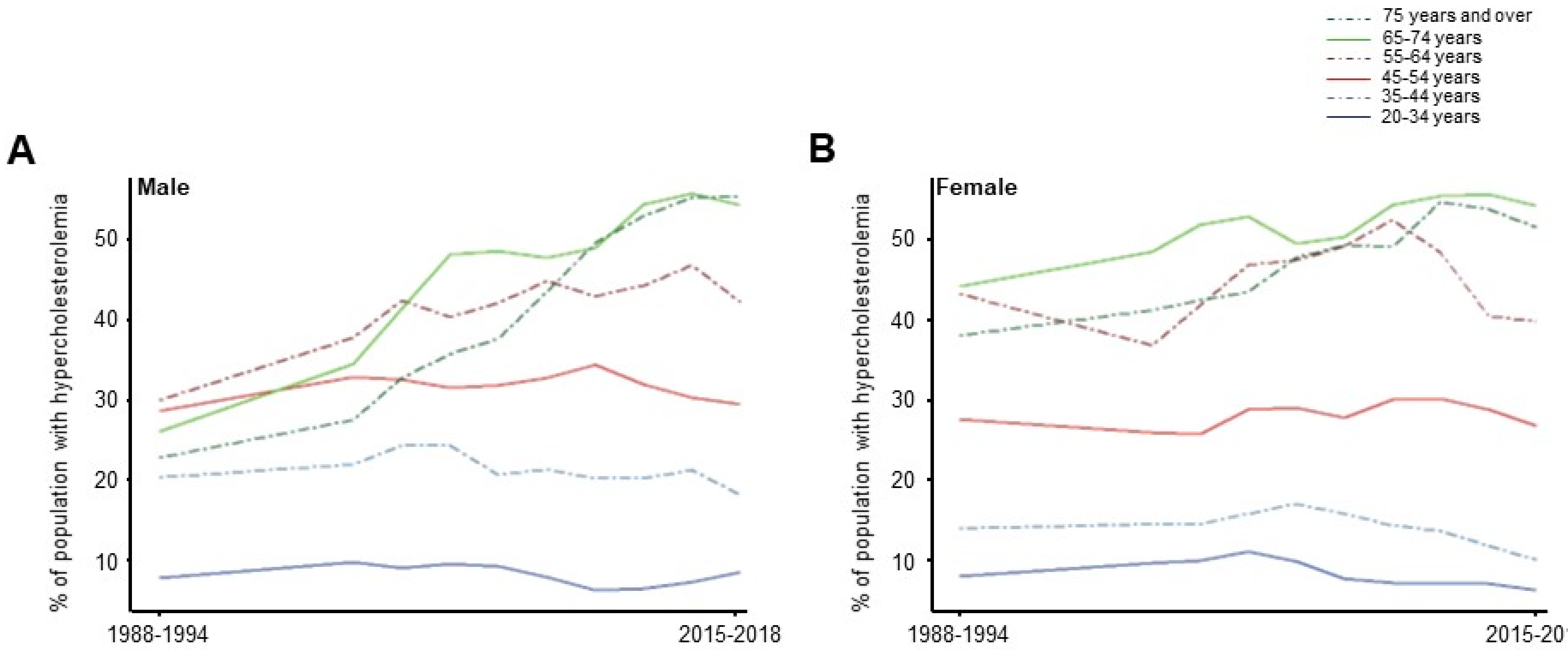


**Supplementary Figure 3. Assessment of population percentage with hypercholesterolemia according to age.**

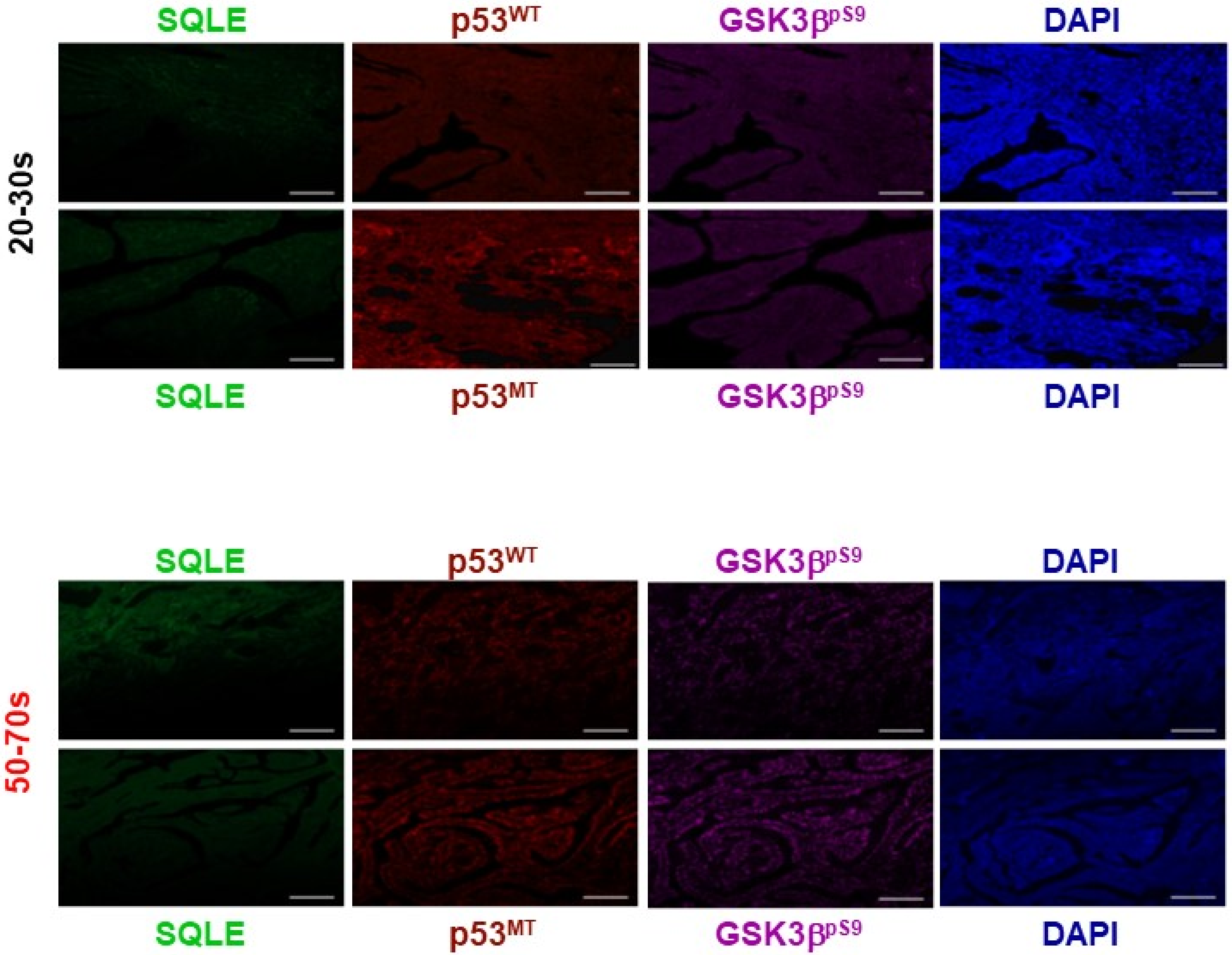


**Supplementary Figure 4. The expression levels of the candidates in thyroid cancers**

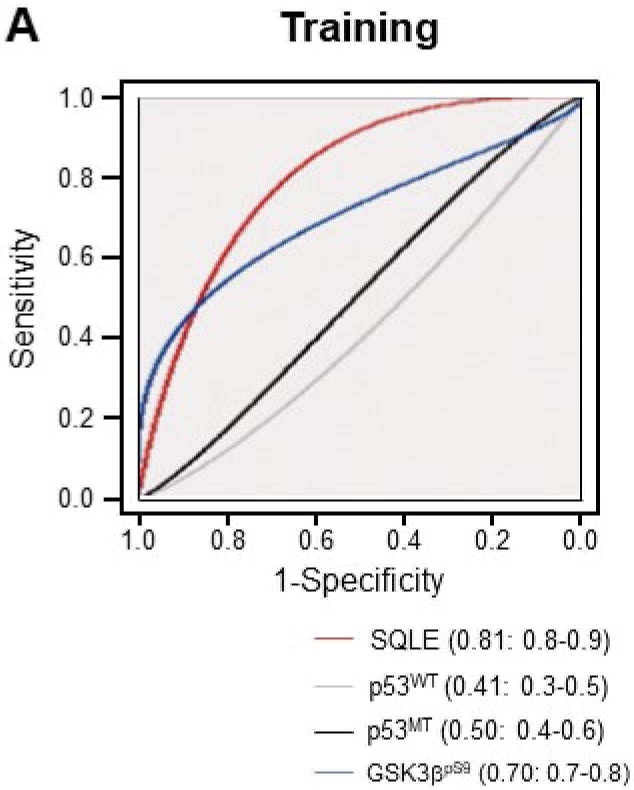


**Supplementary Figure 5. The ROC curves for diagnosing CRCs using levels of each candidate**
