## Supplementary Table 1 for "Reducing squalene epoxidase by the aging-dependent intra-tissue cholesterol accumulation is associated with increased colorectal cancer patient severity in high-risk populations"

**Supplementary Table 1.** Clinicopathological information of a training set generated from the discovery cohort by applying random forest and univariate analysis of variables associated with the survival of CRC patients.

| Training | | | | |
|---|---|---|---|---|
| **Number of all patients** | **< 50 (n = 795)** | | **> 50 (n = 351)** | |
| **Age** | | | | |
| **Patients with cancer** | | | | |
| Mean | 43.8 | | 65.7 | |
| Median ± [a]SD | 45.0 ± 4.8 | | 68.0 ± 8.0 | |
| Range (min - max) | (31 - 48) | | (50 - 86) | |
| **Control subjects** | | | | |
| Mean | 39.1 | | 68.1 | |
| Median ± SD | 40.0 ± 3.2 | | 68.0 ± 4.9 | |
| Range (min - max) | (31 - 45) | | (54 - 81) | |
| **Sex** | **[b]n (%)** | **HR (95% CI), *P* value** | **HR (95% CI), *P* value** | **n (%)** |
| **Patients with cancer** | | | | |
| Male | 371 (22.65) | 0.97 (0.75 - 1.25), 0.83 | 3.12 (1.89 - 5.15), 7x10$^{-6}$ | 219 (13.37) |
| Female | 267 (16.30) | ref. | ref. | 59 (3.60) |
| **Control subjects** | | | | |
| Male | 88 (5.37) | - | - | 51 (3.11) |
| Female | 69 (4.21) | - | - | 22 (1.34) |
| **Subtype** | **n (%)** | **HR (95% CI), *P* value** | **HR (95% CI), *P* value** | **n (%)** |
| **Patients with cancer** | | | | |
| Adenocarcinoma | 537 (32.78) | 0.70 (0.55 - 0.90), 5x10$^{-3}$ | 1.46 (1.06 - 2.00), 0.01 | 262 (16.00) |
| Mucinous adenocarcinoma | 100 (6.11) | 0.11 (0.07 - 0.19), 2x10$^{-16}$ | 0.88 (0.35 - 2.23), 0.79 | 11 (0.67) |
| Papillary carcinoma | - | - | 1.25x10$^{-7}$ (0.00 - inf), 0.99 | 5 (0.31) |
| Signet-ring cell carcinoma | 1 (0.06) | 1.35x10$^{-6}$ (0.00 - inf), 0.98 | - | - |
| **Control subjects** | 157 (9.58) | ref. | ref. | 73 (4.46) |
| **AJCC stage** | **n (%)** | **HR (95% CI), *P* value** | **HR (95% CI), *P* value** | **n (%)** |
| **Patients with cancer** | | | | |
| Stage 1-2 | 204 (12.45) | 0.83 (0.54 - 1.29), 0.42 | 1.87 (1.33 - 2.63), 2x10$^{-4}$ | 144 (8.79) |
| Stage 3-4 | 425 (25.95) | 0.84 (0.56 - 1.26), 0.40 | 1.81 (1.17 - 2.78), 6x10$^{-3}$ | 115 (7.02) |
| unknown | 9 (0.56) | - | - | 19 (1.16) |
| **Control subjects** | 157 (9.58) | ref. | ref. | 73 (4.46) |
| **The TNM staging** | **n (%)** | **HR (95% CI), *P* value** | **HR (95% CI), *P* value** | **n (%)** |

| | | | | |
|---|---|---|---|---|
| **Patients with cancer** | | | | |
| T3N0M0 (T1N0M0, T2N0M0) | 101 (6.17) | 0.60 (0.39 - 0.93), 0.02 | 1.09 (0.70 - 1.70), 0.68 | 61 (3.72) |
| T3N1M0 (T3N1M1) | 160 (9.77) | 0.92 (0.66 - 1.30), 0.67 | 1.21 (0.65 - 2.26), 0.53 | 37 (2.26) |
| T3N2M0 (T3N2M1) | 119 (7.27) | 0.99 (0.68 - 1.44), 0.97 | 2.23 (0.94 - 5.29), 0.06 | 15 (0.92) |
| T4N0M0 (T4N0M1) | 71 (4.34) | 0.28 (0.19 - 0.43), $3.7 \times 10^{-9}$ | 2.18 (1.44 - 3.30), $2 \times 10^{-4}$ | 79 (4.82) |
| T4N1M0 (T4N1M1) | 87 (5.31) | 1.02 (0.73 - 1.44), 0.86 | 1.08 (0.57 - 2.04), 0.80 | 16 (0.98) |
| T4N2M0 (T4N2M1) | 52 (3.17) | 1.03 (0.69 - 1.55), 0.85 | 1.29 (0.86 - 1.93), 0.21 | 50 (3.05) |
| unknown | 48 (2.93) | - | - | 20 (1.22) |
| **Control subjects** | 157 (9.58) | ref. | ref. | 73 (4.46) |
| **Tumour differentiation** | **n (%)** | **HR (95% CI), *P* value** | **HR (95% CI), *P* value** | **n (%)** |
| **Patients with cancer** | | | | |
| G1 | 186 (11.36) | 0.83 (0.59 - 1.17), 0.30 | 1.38 (0.94 - 2.01), 0.09 | 86 (5.25) |
| G2 | 343 (20.94) | 0.82 (0.63 - 1.08), 0.17 | 1.42 (0.99 - 2.05), 0.05 | 121 (7.39) |
| G3 | 63 (3.85) | 1.04 (0.71 - 1.53), 0.81 | 1.30 (0.77 - 2.19), 0.31 | 44 (2.69) |
| unknown | 46 (2.81) | - | - | 27 (1.64) |
| **Control subjects** | 157 (9.58) | ref. | ref. | 73 (4.46) |
| **(c)Follow-up** | | **HR (95% CI), *P* value** | **HR (95% CI), *P* value** | |
| **Patients with cancer** | | 0.99 (0.98 - 0.99), $2.26 \times 10^{-14}$ | 1.02 (1.01 - 1.02), $< 2 \times 10^{-16}$ | |
| Mean | 89.4 | | | 101 |
| Median ± SD | 122.0 ± 47.0 | | | 99.0 ± 29.4 |
| Range (min-max) | (2 - 100) | | | (40 - 140) |
| **Control subjects** | | ref. | ref. | |
| Mean | 95 | | | 110 |
| Median ± SD | 124.0 ± 42.0 | | | 123.0 ± 31.0 |
| Range (min-max) | (30 - 100) | | | (54 - 137) |
| **Candidates** | **(Median ± SD)** | **HR (95% CI), *P* value** | **HR (95% CI), *P* value** | **(Median ± SD)** |
| (d)$p53^{WT}$ | | | | |
| **Patients with cancer** | 0.30 ± 5.44 | 1.00 (0.98 - 1.02), 0.41 | 1.33 ($7.4 \times 10^{-3}$ - $2.4 \times 10^{2}$), 0.91 | -0.1 ± 0.00 (*P* = 0.02) |
| **Control subjects** | 0.003 ± 0.01 | | | -0.001 ± 0.01 |
| (e)$p53^{MT}$ | | | | |
| **Patients with cancer** | 0.32 ± 0.27 | 1.44 (1.12 - 1.83), $3 \times 10^{-3}$ | 1.38 (0.73 - 2.62), 0.31 | 0.29 ± 0.20 (*P* = 0.03) |
| **Control subjects** | 0.35 ± 0.27 | | | 0.35 ± 0.25 |
| (f)$GSK3\beta^{pS9}$ | | | | |
| **Patients with cancer** | 0.24 ± 0.14 | 1.03 (1.01 - 1.05), $3 \times 10^{-3}$ | 1.30 (0.89 - 1.90), 0.16 | 0.34 ± 0.15 (*P* = $1 \times 10^{-8}$) |
| **Control subjects** | 0.99 ± 1.00 | | | 0.02 ± 1.15 |

| SQLE | | | | |
|---|---|---|---|---|
| **Patients with cancer** | 0.42 ± 12.04 | 0.98 (0.96 - 1.01), 0.36 | 2.03 (1.27 - 3.23), $2x10^{-3}$ | 0.21 ± 0.30 (*P* = 0.02) |
| **Control subjects** | 0.01 ± 0.07 | | | 0.41 ± 0.71 |

**(a)** SD: standard deviation

**(b)** n (%)**:** Calculated in the following way: %= (n/1638) *100

**(c)** Follow-up: the period (in months) from diagnosis to an event (death, survival, or loss of contact)

**(d)** $p53^{WT}$: mainly detecting wild-type p53 (DO-1: SC-126, SCBT)

**(e)** $p53^{MT}$: to detect mutant p53 (Y5: ab32049, Abcam)

**(f)** $GSK3\beta^{pS9}$: the inactive form of GSK3β (the anti-$GSK3\beta^{pS9}$ antibody; 9323s, Cell Signaling Technology)
