## Supplementary Table 2 for "Reducing squalene epoxidase by the aging-dependent intra-tissue cholesterol accumulation is associated with increased colorectal cancer patient severity in high-risk populations"

**Supplementary Table 2.** Clinicopathological information of the testing set generated from the discovery cohort by applying random forest and univariate analysis of variables associated with the survival of CRC patients.

| **Testing** | | | | |
|---|---|---|---|---|
| **Number of all patients** | **< 50 (n = 357)** | | **> 50 (n = 135)** | |
| **Age** | | | | |
| **Patients with cancer** | | | | |
| Mean | 42.1 | | 66.2 | |
| Median ± [a]SD | 42.0 ± 4.4 | | 68.0 ± 7.4 | |
| Range (min - max) | (35 - 49) | | (50 - 82) | |
| **Control subjects** | | | | |
| Mean | 37.7 | | 68.3 | |
| Median ± SD | 30.0 ± 4.4 | | 68.0 ± 0.4 | |
| Range (min - max) | (25 - 47) | | (52 - 82) | |
| **Sex** | **[b]n (%)** | **HR (95% CI), *P* value** | **HR (95% CI), *P* value** | **n (%)** |
| **Patients with cancer** | | | | |
| Male | 163 (9.95) | 1.08 (0.70 - 1.68), 0.70 | 3.15 (1.37 - 7.23), $6x10^{-3}$ | 75 (4.58) |
| Female | 125 (7.63) | ref. | ref. | 32 (1.95) |
| **Control subjects** | | | | |
| Male | 43 (2.63) | - | - | 19 (1.16) |
| Female | 26 (1.59) | - | - | 9 (0.56) |
| **Subtype** | **n (%)** | **HR (95% CI), *P* value** | **HR (95% CI), *P* value** | **n (%)** |
| **Patients with cancer** | | | | |
| Adenocarcinoma | 257 (15.69) | 0.82 (0.58 - 1.16), 0.28 | 1.43 (0.85 - 2.38), 0.16 | 100 (6.11) |
| Mucinous adenocarcinoma | 31 (1.89) | 0.27 (0.14 - 0.52), $7x10^{-5}$ | 4.14 (0.94 - 18.13), 0.05 | 3 (0.18) |
| Papillary carcinoma | - | - | $4.78x10^{-8}$ (0.00 - inf), 0.99 | 4 (0.24) |
| Signet-ring cell carcinoma | - | - | - | - |
| **Control subjects** | 69 (4.21) | ref. | ref. | 28 (1.71) |
| **AJCC stage** | **n (%)** | **HR (95% CI), *P* value** | **HR (95% CI), *P* value** | **n (%)** |
| **Patients with cancer** | | | | |
| Stage 1-2 | 75 (4.58) | 0.80 (0.49 - 1.30), 0.38 | 1.86 (1.06 - 3.27), 0.03 | 48 (2.93) |
| Stage 3-4 | 211 (12.88) | 0.91 (0. 61- 1.36), 0.66 | 1.86 (1.05 - 3.29), 0.03 | 49 (2.99) |
| unknown | 2 (0.12) | - | - | 10 (0.61) |
| **Control subjects** | 69 (4.21) | ref. | ref. | 28 (1.71) |
| **The TNM staging** | **n (%)** | **HR (95% CI), *P* value** | **HR (95% CI), *P* value** | **n (%)** |

| | | | | |
|---|---|---|---|---|
| **Patients with cancer** | | | | |
| T3N0M0 (T1N0M0, T2N0M0) | 39 (2.38) | 0.93 (0.44 - 1.98), 0.86 | 1.09 (0.45 - 2.66), 0.83 | 12 (0.73) |
| T3N1M0 (T3N1M1) | 79 (4.82) | 0.91 (0.53 - 1.54), 0.73 | 1.30 (0.53 - 3.16), 0.55 | 19 (1.16) |
| T3N2M0 (T3N2M1) | 55 (3.36) | 0.94 (0.50 - 1.75), 0.85 | 1.56 (0.51 - 4.69), 0.42 | 4 (0.24) |
| T4N0M0 (T4N0M1) | 34 (2.08) | 0.45 (0.27 - 0.75), $2x10^{-3}$ | 2.15 (1.19 - 3.87), 0.01 | 34 (2.08) |
| T4N1M0 (T4N1M1) | 37 (2.26) | 0.68 (0.40 - 1.15), 0.15 | 1.19 (0.15 - 9.03), 0.86 | 5 (0.31) |
| T4N2M0 (T4N2M1) | 33 (2.01) | 0.96 (0.57 - 1.61), 0.89 | 1.37 (0.72 - 2.59), 0.33 | 23 (1.40) |
| unknown | 11 (0.67) | - | - | 10 (0.61) |
| **Control subjects** | 69 (4.21) | ref. | ref. | 28 (1.71) |
| **Tumour differentiation** | **n (%)** | **HR (95% CI), *P* value** | **HR (95% CI), *P* value** | **n (%)** |
| **Patients with cancer** | | | | |
| G1 | 86 (5.25) | 0.82 (0.52 - 1.31), 0.41 | 1.26 (0.69 - 2.31), 0.45 | 38 (2.32) |
| G2 | 136 (8.30) | 0.81 (0.55 - 1.20), 0.31 | 1.26 (0.70 - 2.27), 0.42 | 46 (2.81) |
| G3 | 41 (2.50) | 1.07 (0.62 - 1.83), 0.79 | 1.60 (0.77 - 3.30), 0.20 | 14 (0.85) |
| unknown | 25 (1.53) | - | - | 9 (0.55) |
| **Control subjects** | 69 (4.21) | ref. | ref. | 28 (1.71) |
| **[c]Follow-up** | | **HR (95% CI), *P* value** | **HR (95% CI), *P* value** | |
| **Patients with cancer** | | 0.99 (0.98 - 0.99), $2x10^{-8}$ | 1.02 (1.01 - 1.03), $5x10^{-9}$ | |
| Mean | 85.4 | | | 100 |
| Median ± SD | 110.0 ± 48.4 | | | 100.0 ± 20.1 |
| Range (min-max) | (2 - 100) | | | (40 - 140) |
| **Control subjects** | | ref. | ref. | |
| Mean | 103.4 | | | 113 |
| Median ± SD | 124.0 ± 43.9 | | | 114.0 ± 27.3 |
| Range (min-max) | (30 - 100) | | | (54 - 137) |
| **Candidates** | **(Median ± SD)** | **HR (95% CI), *P* value** | **HR (95% CI), *P* value** | **(Median ± SD)** |
| [d]$p53^{WT}$ | | | | |
| **Patients with cancer** | 0.58 ± 0.09 | 1.01 (0.97 - 1.05), 0.57 | 0.41 ($6.7x10^{-4}$ - 255), 0.78 | 0.01 ± 0.02 (*P* = 0.05) |
| **Control subjects** | 0.001 ± 0.009 | | | -0.007 ± 0.003 |
| [e]$p53^{MT}$ | | | | |
| **Patients with cancer** | 0.55 ± 0.20 | 1.46 (1.07 - 1.98), 0.01 | 1.30 (0.48 - 3.49), 0.59 | 0.40 ± 0.13 (*P* = 0.01) |
| **Control subjects** | 0.43 ± 0.25 | | | 0.32 ± 0.25 |
| [f]$GSK3\beta^{pS9}$ | | | | |
| **Patients with cancer** | 0.16 ± 0.16 | 1.04 (1.01 - 1.08), $6x10^{-3}$ | 1.29 (0.58 - 2.86), 0.53 | 0.25 ± 0.07 (*P* = 3 x $10^{-3}$) |
| **Control subjects** | 0.09 ± 0.82 | | | 0.53 ± 0.03 |

| | | | | |
|---|---|---|---|---|
| SQLE | | | | |
| **Patients with cancer** | 0.52 ± 3.24 | 0.99 (0.92 - 1.06), 0.80 | 1.92 (1.00 - 3.71), 0.04 | 0.10 ± 0.24 (*P* = 0.01) |
| **Control subjects** | 0.09 ± 0.55 | | | 0.40±0.52 |

**(a)** SD: standard deviation

**(b)** n (%)**:** Calculated in the following way: %= (n/1638) *100

**(c)** Follow-up: the duration (in months) of an event (death, survival, or loss of contact) was observed since a diagnosis was made

**(d)** $p53^{WT}$: mainly detecting wild-type p53 (DO-1: SC-126, SCBT)

**(e)** $p53^{MT}$: to detect mutant p53 (Y5: ab32049, Abcam)

**(f)** $GSK3\beta^{pS9}$: the inactive form of GSK3β (the anti-$GSK3\beta^{pS9}$ antibody; 9323s, Cell Signaling Technology)
